# Traumatic Encephalopathy Syndrome in the Late Effects of Traumatic Brain Injury (LETBI) study cohort

**DOI:** 10.1101/2024.07.04.24309955

**Authors:** Kristen Dams-O’Connor, Enna Selmanovic, Ariel Pruyser, Lisa Spielman, Ashlyn Bulas, Eric Watson, Jesse Mez, Jeanne M. Hoffman

## Abstract

**Background and Objective:** Traumatic encephalopathy syndrome (TES) is the proposed clinical manifestation of chronic traumatic encephalopathy (CTE) neuropathologic change secondary to repetitive head impacts (RHI). The prevalence of TES and its component symptoms is not known in individuals with single TBI, a subset of whom also have RHI. We used prospectively collected data to operationalize TES criteria and test the hypothesis that the core clinical features of TES are common among those with TBI, regardless of RHI exposure status and other demographic and injury characteristics.

**Methods:** Secondary analysis of data from the Late Effects of TBI (LETBI) study, a community-based study of individuals with complicated mild, moderate, or severe TBI. Participants were categorized by TBI severity and presence of RHI, creating 6 groups (those with isolated mild, moderate, and severe TBI, with and without RHI). Chi-squared tests were used to compare the proportion of each group that met each of the core clinical criteria overall TES diagnosis. Binary logistic regression models were used to examine associations of demographic and injury characteristics on TES diagnosis.

**Results:** In 295 participants with TBI, mean (SD) age 52.6(15.6) years and 35.6% female, 138 (46.8%) had RHI exposure meeting the TES criteria exposure threshold. In the full sample, 56.9%, 32.9% and 45.8% of participants met TES core criterion of cognitive impairment, neurobehavioral dysregulation, and progressive course of clinical features, respectively. Overall, 14.9% of the LETBI sample had substantial RHI exposure and met all 3 clinical features, meeting consensus-based TES criteria. When RHI exposure criterion was lifted, 33.5% of the LETBI sample with isolated TBI met all core clinical criteria. No injury or demographic variables predicted the likelihood of meeting TES Core Criteria (OR=3.02, p=0.10).

**Discussion:** Rates of TES clinical features are high among TBI survivors with and without RHI, across injury severity groups. Presence of TES core clinical features was greatest among those with no RHI, suggesting that chronic and sometimes progressive clinical sequelae of TBI resemble TES, but may reflect a distinct pathobiological process. Limitations include possible selection of participants with chronic symptoms. Findings emphasize the centrality of RHI exposure to TES diagnostic criteria.

## INTRODUCTION

Early clinical observations of adverse effects of exposure to repetitive head impacts (RHI) in boxers, ^1,2^ seen in some cases following a single traumatic brain injury (TBI), ^3^ are often cited in contemporary studies of postmortem chronic traumatic encephalopathy (CTE) neuropathology in American football players, ^4^ implying that the same underlying disease process (and clinical manifestations thereof) is being described.

Consensus-based definitions of CTE neuropathological criteria have now been refined^5^ to better distinguish CTE from other neurodegenerative and age-related processes by identifying pathognomonic lesion(s). CTE is regarded as a progressive neurodegenerative disease resulting from prolonged exposure to RHI. Extant studies have primarily included male American-style football players, and far less is known about CTE or its potential clinical correlates in other RHI- and TBI-exposed populations.

Parallel efforts have endeavored to characterize the constellation of clinical symptoms described in individuals who have been posthumously diagnosed with CTE^6-9^ to permit in-vivo diagnosis. Research diagnostic criteria for traumatic encephalopathy syndrome (TES), the proposed clinical manifestation of CTE neuropathology, were described in 2014^9^ based on symptoms observed during life in 202 cases with CTE neuropathology. Central features included impairments in cognition, behavior, and mood. ^9^ Concerns about over-diagnosing TES per these broad criteria are articulated in a series of papers^10-12^ that reported 65-83% of men with intermittent explosive disorder or a recent major depressive episode (respectively) met TES criteria. ^11^ A central limitation of these studies, ^10-12^ was the unavailability of data on lifetime exposure to head trauma, which effectively prevents application of the foundational exposure criteria to the dataset.

In 2019, the National Institute of Neurological Disorders and Stroke (NINDS) convened a Consensus Workshop to define in vivo TES diagnostic criteria. ^13^ The consensus panel identified core clinical criteria for TES as cognitive impairment and/or neurobehavioral dysregulation, and progressive course of decline. Throughout four rounds of a modified Delphi procedure in the workshop, the panel ultimately decided that isolated moderate-severe TBI, regardless of number of isolated TBIs, did not meet the exposure criterion, largely due to the paucity of research on the prevalence of CTE among individuals with single moderate-severe TBI. ^13-15^ Substantial overlap in the clinical presentation of individuals with CTE and many hallmark clinical sequelae of isolated TBI^16-20^ may pose challenges to in-vivo diagnosis of the clinical correlate of CTE, and the panel emphasized the need for refinement of TES criteria in cohorts with diverse patterns of head trauma exposure.

In the current paper, we apply the consensus-based TES research diagnostic criteria to data gathered from the Late Effects of TBI (LETBI) study cohort of individuals with TBI, many of whom also have RHI exposure. We test the hypothesis that the core clinical features of TES are common among those with TBI, regardless of RHI exposure status. The goals of this work are to illustrate the process of operationalizing TES criteria (developed based on qualitative information provided by loved ones of decedents who were diagnosed with CTE) using prospectively collected in-vivo data, and to investigate whether any demographic or injury factors predicted likelihood of TES diagnosis.

## METHODS

### Study design and population

Participants in this study were enrolled in the Late Effects of Traumatic Brain Injury (LETBI) project, an ongoing multicenter prospective longitudinal brain donor program designed to identify the clinical characteristics and postmortem neuropathology of post-traumatic neurodegeneration. ^21^ Participants are recruited from TBI research registries at two sites and from community-based settings. Research registries included individuals who had participated in prior research projects and indicated a willingness to be contacted for future studies, as well as TBI survivors and care providers who signed up to receive newsletters and informational resources about brain injury. Participants were additionally recruited from the community through TBI screening initiatives and informational flyers providing information about TBI and the research project. Eligible participants met the following criteria: (1) per comprehensive structured interview and medical record review were determined to have sustained at least one complicated mild TBI (defined as a blow to the head resulting in a period of altered mental status ≤24 hours and/or unconsciousness ≤30 minutes, ^22^ but with intracranial findings detected on acute neuroimaging), ^23,24^ moderate TBI, severe TBI, OR ≥2 mild TBIs, (2) are at least 1 year post first head trauma exposure, (3) aged ≥18 years, and (4) are English speaking. ^25^

### Standard Protocol Approvals, Registrations and Patient Consents

Study procedures were approved by the Icahn School of Medicine at Mount Sinai’s Program for the Protection of Human Subjects/Institutional Review Board prior to data collection. All participants or their legally authorized representatives provided written informed consent prior to participation.

### Head trauma exposure ascertainment

Lifetime history of TBI and/or RHI was characterized using the Brain Injury Screening Questionnaire (BISQ), a structured interview for lifetime head trauma exposure. ^26^ The BISQ queries blows to the head with 20 contextual recall cues (e.g., in a vehicular accident, on the playground, etc.). For each reported event, subsequent items query the presence and duration of altered mental status and/or unconsciousness, permitting injury severity classification per standard criteria. ^27^ Additional modules characterize duration and age(s) of exposure to RHI sustained in the context of organized contact sports, military service, and intimate partner violence (IPV). For all RHI etiologies, age at start/stop of exposure, and overall duration of exposure is queried. Head trauma exposure history was verified through review of medical records whenever available; neuroimaging records were also used to determine the presence of intracranial lesions which informed TBI severity classification (e.g., complicated vs. uncomplicated mild TBI). Use of these data to determine exposure thresholds for TES criteria is detailed in Supplementary Table S1.

### LETBI Neurobehavioral Assessment Battery

All participants completed a comprehensive standardized evaluation of cognitive function across multiple domains, behavior, mood, and overall health and function; the LETBI battery is primarily comprised of NIH Common Data Elements (CDEs^28^; See Table S1) and additional measures selected to overlap with large-scale dementia studies. ^29-31^

### TES Criteria

We used all available data to operationalize diagnostic criteria for TES, ^13^ as described in Table S1. Individuals meeting TES Diagnostic Criteria must have: (1) substantial exposure to RHI; (2) core clinical features of cognitive impairment and/or neurobehavioral dysregulation, AND evidence of a worsening/progressive clinical course; and (3) clinical symptoms cannot be fully accounted for by another disease. Decisions regarding the operational definition of TES criteria were made by expert consensus (KDOC, EW, JH, JM) based on the measures available in the LETBI study and previous literature when applicable.

#### Substantive Exposure to Repetitive Head Impacts

We used BISQ data to define RHI exposure (Criterion 1) per Katz et al., including RHI etiologies that are very prescriptive (e.g., ≥5 years of American Football, ≥2 of which at the high school level or beyond), as well as those that are more exploratory, e.g., “exposure to multiple blast and other explosions,” and “exposure to multiple head impacts over an extended period of time” (see Table S1).

#### Core Clinical Features

As the Katz TES criteria are based primarily on the retrospective report of family members whose loved ones have been diagnosed with CTE neuropathology, defining the core clinical features (Criterion 2) ^13^ with psychometric data collected during life requires clinical judgment. The criteria acknowledge that performance-based data are not always available; as detailed in Table S1, we used a combination of self- and informant-report measures alongside formal neuropsychological test results when available.

Katz et al^13^ require that 4 indicators must be present to meet criteria for the first Core Clinical Feature, Cognitive Impairment: self- or informant/clinician-reported cognitive impairment, decline from baseline function, deficits in episodic memory and/or executive function, and impaired performance on neuropsychological testing (when available). We modified a demographic-based algorithm^32^ to estimate premorbid functioning based on age and education. ^13,33,34^ The TES criteria define “substantially impaired performance” on neuropsychological tests as 1.5 standard deviations (SD) below “appropriate norms, accounting for the individual’s estimated premorbid functioning;” we used normative data to calculate standard scores on each measure as detailed in

Table S1. Absent clear criteria to define impairment, we used 1.5 SD below normative groups to define domain-specific deficits and global impairment per performance-based tests.

Neurobehavioral dysregulation requires that the following criteria are met: self- or informant/clinician-reported neurobehavioral dysregulation, changes from baseline function, significant and poor regulation/control of emotions and behavior (as defined with examples provided by Katz et al^13^). We used empirically validated cut-points on psychometric scales when possible, in addition to endorsement of symptoms/behaviors provided as examples in the TES criterion definition (see Table S1).

To operationalize progressive worsening of clinical features, we used objective performance-based data for participants with ≥2 study visits (see Table S1). Absent detailed guidance, we evaluated an operational definition of decline as 1.5 SD worsening of scores between visits. We used measures that specifically asked participants to compare current function to their highest achieved level of post-injury function to distinguish enduring but stable deficits (which might allow an individual to meet criteria for cognitive impairment and/or neurobehavioral dysregulation) from post-recovery decline (i.e., progressive course).

#### Not Fully Accounted for by Other Disorders

The presence of a neurodegenerative, medical, psychiatric, or other condition that “fully” accounts for the core clinical features is exclusionary for a TES diagnosis, but diagnosis of conditions do not otherwise exclude a TES diagnosis (Criterion 3). ^13^ Determination of whether a given condition fully accounts for all symptoms requires case-by-case evaluation. As such, and in the spirit of hypothesis-generation, we present the prevalence of known co-occurring conditions in those with and without RHI who do and do not meet the core clinical TES criteria.

#### Level of Functional Dependence/Dementia

TES criterion 4 outlines five levels of function ranging from to “Severe” to “Independent.” We triangulated ratings from validated assessments of cognitive and social functioning and activities of daily living (see Table S1), and assigned the level of functional dependence corresponding to the lowest (most impaired) reported function across available measures.

### Provisional Levels of Certainty for CTE Pathology

Katz et al^13^ suggest criteria to facilitate exploration that TES features reflect an underlying diagnosis of CTE. We used available data to operationalize supportive features of likely neuropathology that go beyond TES (i.e. intensity of RHI exposure, functional independence, motor signs, and psychiatric features) ^13^ as outlined in Supplementary Table S2.

### Statistical Analysis

Data analyses were completed in SAS 9.4^35^ and IBM SPSS 29.0. ^36^ We categorized the sample by TBI severity (mild/moderate/severe) and presence of RHI history, thereby forming 6 groups (i.e., those with isolated TBI of each severity, with and without RHI). If a person reported more than one isolated TBI, we grouped them based on their most severe injury. We calculated the proportion of each group that met each of the core clinical criteria; we also examined level of functional dependence and levels of certainty (for CTE neuropathology). ^13^ We explored the proportion of the sample meeting TES criteria if the RHI exposure criteria were lifted to better understand the prevalence of TES clinical criteria in those with isolated TBI. We compared the proportion of the sample who met each of the core clinical criteria and overall TES diagnosis between those with vs. without RHI exposure using chi-squared tests, and calculated the rates of medical and behavioral health conditions among the TBI-RHI groups. We used binary logistic regression models to examine associations of demographic and injury characteristics on TES diagnosis.

### Data Availability

Deidentified data sets included herein may be shared at the request of any qualified investigator for the purposes of replicating procedures and results. All data requests can be made through the National Institute of Health’s Federal Interagency Traumatic Brain Injury Research Informatics System (FITBIR).

## RESULTS

### Sample characteristics

A total of 417 participants were enrolled in the LETBI study as of March 2024, and 295 had complete data on key variables (Figure 1). The mean (SD) age at baseline LETBI visit was 52.6 (15.6) years, mean age at most severe injury was 38.6 (18.8) years, and the sample was 85.1% White, 15.9% Hispanic or Latino, and 64.4% male. Sample demographics are presented by injury exposure history in Table 1, and RHI etiologies are presented in Supplementary Table S3.

**Table 1.** Sample characteristics by head trauma exposure group.

| LETBI Demographics |  | ≥2 Mild TBIs Only<br>N=35 | ≥2 Mild TBIs + RHI<br>N=43 | Complicated Mild/Moderate Only<br>N=32 | Complicated Mild/Moderate + RHI<br>N=23 | Severe Only<br>N=90 | Severe + RHI<br>N=72 | Total<br>N=295 |
| --- | --- | --- | --- | --- | --- | --- | --- | --- |
| **Age** |  |  |  |  |  |  |  |  |
|  | Mean ± SD range | 52.43±13.7<br>28-74 | 45.37±16.27<br>21-74 | 59.87±16.51<br>23-89 | 57.7±15.08<br>26-80 | 55.4±13.8 22-83 | 48.6±15.6<br>22-80 | 52.6±15.5<br>21-89 |
| **Education Years** |  |  |  |  |  |  |  |  |
|  | Mean ± SD | 15.43±4.13 | 16.31±4.79 | 15.71±4.21 | 15.68±3.34 | 15.2±3.88 | 14.55±3.04 | 15.31±3.8 |
| **Age at Injury** |  |  |  |  |  |  |  |  |
|  | Mean ± SD | 40.77±20.23 | 27.52±16.87 | 47.9±22.72 | 39.29±17.74 | 40.31±18.19 | 37.06±15.69 | 38.56±18.8 |
| **Time Since Injury** |  |  |  |  |  |  |  |  |
|  | Mean ± SD | 11.27±11.01 | 18.43±16.74 | 12.54±10.65 | 19.96±16.27 | 15.58±9.74 | 11.48±6.97 | 14.53±11.5 |
| **Number of TBIs** |  |  |  |  |  |  |  |  |
|  | Mean ± SD | 1.47±1.5 | 2.62±3.13 | 1.55±1.08 | 2.59±1.79 | 1.44±0.95 | 2.77±2.98 | 2.02±2.06 |
| **Time Between Visits** |  |  |  |  |  |  |  |  |
|  | Mean ± SD | 3.05±1.13 | 3.81±2.08 | 5.53±1.81 | 5.10±2.57 | 4.71±1.61 | 4.10±2.00 | 4.52±1.93 |
| **Male** |  |  |  |  |  |  |  |  |
|  | N, (%) | 16(45.7%) | 31(72.1%) | 21(65.6%) | 15(65.2%) | 53(58.9%) | 54(75%) | 190(64.4%) |
| **Race** |  |  |  |  |  |  |  |  |
|  | American Native | 0(0%) | 0(0%) | 0(0%) | 1(4.3%) | 2(2.2%) | 0(0%) | 3(1%) |
|  | Asian | 0(0%) | 0(0%) | 1(3.1%) | 0(0%) | 3(3.3%) | 0(0%) | 4(1.4%) |
|  | African American | 2(5.7%) | 3(7%) | 3(9.4%) | 0(0%) | 3(3.3%) | 2(2.8%) | 13(4.4%) |
|  | Pacific Islander | 0(0%) | 0(0%) | 0(0%) | 0(0%) | 0(0%) | 2(2.8%) | 2(0.7%) |
|  | White | 30(85.7%) | 39(90.7%) | 27(84.4%) | 21(91.3%) | 73(81.1%) | 61(84.7%) | 251(85.1%) |
|  | Other | 0(0%) | 0(0%) | 1(3.1%) | 0(0%) | 4(4.4%) | 3(4.2%) | 8(2.7%) |
|  | [Missing] | 3(8.6%) | 1(2.3%) | 0(0%) | 1(4.3%) | 5(5.6%) | 4(5.6%) | 14(4.7%) |
| **Hispanic/Latino** |  |  |  |  |  |  |  |  |
|  | N, (%) | 7(20%) | 5(11.6%) | 2(6.3%) | 3(13%) | 18(20.0%) | 12(16.7%) | 47(15.9%) |
|  | [Missing] | 3(8.6%) | 2(4.7%) | 3(9.4%) | 4(17.4%) | 10(11.1%) | 6(8.3%) | 28(9.5%) |
| **Marital Status** |  |  |  |  |  |  |  |  |
|  | Never Married | 10(28.6%) | 13(30.2%) | 5(15.6%) | 6(26.1%) | 25(27.8%) | 22(30.6%) | 81(27.5%) |
|  | Married | 13(37.1%) | 19(44.2%) | 13(40.6%) | 9(39.1%) | 34(37.6.8%) | 19(26.4%) | 107(36.3%) |
|  | Domestic Partner | 0(0%) | 1(2.3%) | 3(9.4%) | 2(8.7%) | 2(2.2%) | 2(2.8%) | 10(3.4%) |
|  | Separated | 0(0%) | 0(0%) | 1(3.1%) | 1(4.3%) | 2(2.2%) | 3(4.2%) | 7(2.4%) |
|  | Divorced | 6(17.1%) | 7(16.3%) | 8(25%) | 1(4.3%) | 19(21.1%) | 14(19.4%) | 55(18.6%) |
|  | Widowed | 2(5.7%) | 0(0%) | 1(3.1%) | 2(8.7%) | 3(3.3%) | 3(4.2%) | 11(3.7%) |
|  | [Missing] | 4(11.4%) | 3(7.0%) | 1(3.1%) | 2(8.7%) | 5(5.6%) | 9(12.5%) | 24(8.1%) |
| **Employment** |  |  |  |  |  |  |  |  |
|  | Working Now | 14(40%) | 24(55.8%) | 12(37.5%) | 8(34.8%) | 31(34.4%) | 24(33.3%) | 113(38.3%) |
|  | Laid Off | 0(0%) | 0(0%) | 0(0%) | 0(0%) | 1(1.1%) | 0(0%) | 1(0.3%) |
|  | Looking for Work | 1(2.9%) | 0(0%) | 1(3.1%) | 1(4.3%) | 8(8.9%) | 3(4.2%) | 14(4.7%) |
|  | Retired | 7(20%) | 8(18.6%) | 12(37.5%) | 5(21.7%) | 19(21.1%) | 11(15.3%) | 62(21%) |
|  | Disabled | 9(25.7%) | 7(16.3%) | 4(12.5%) | 6(26.1%) | 24(26.7%) | 19(26.4%) | 69(23.4%) |
|  | Keeping House | 1(2.9%) | 0(0%) | 0(0%) | 0(0%) | 1(1.1%) | 2(2.8%) | 4(1.4%) |
|  | Full-Time Student | 0(0%) | 1(2.3%) | 1(3.1%) | 1(4.3%) | 2(2.2%) | 3(4.2%) | 8(2.7%) |
|  | Other | 0(0%) | 0(0%) | 1(3.1%) | 0(0%) | 0(0%) | 0(0%) | 1(0.3%) |
|  | Maternity Leave | 0(0%) | 0(0%) | 0(0%) | 0(0%) | 0(0%) | 1(1.4%) | 1(0.3%) |
|  | [Missing] | 3(8.6%) | 3(7.0%) | 1(3.1%) | 2(8.7%) | 4(4.4%) | 9(12.5%) | 22(7.5%) |

**Figure 1:**
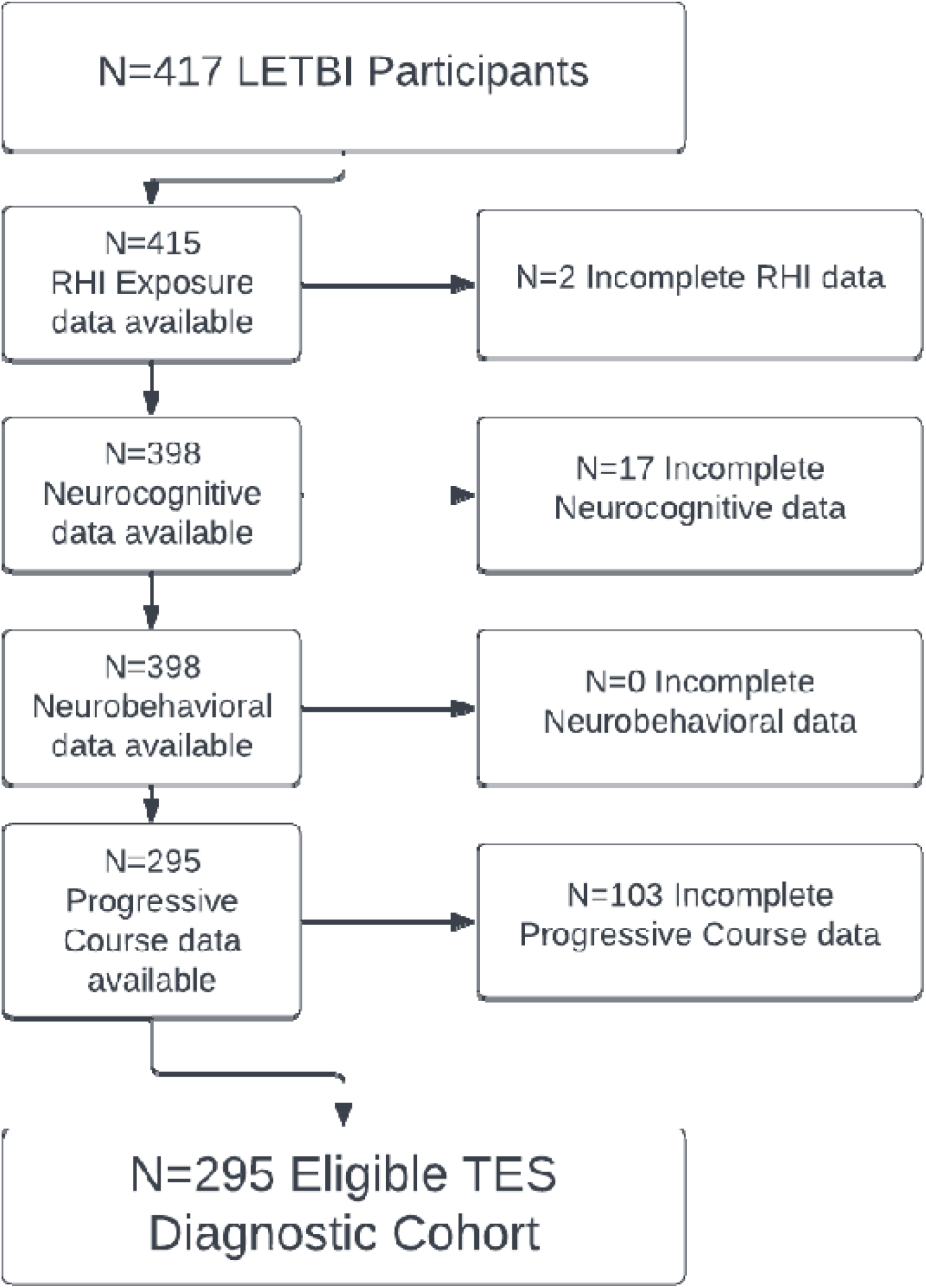
Flow diagram of data availability for TES Diagnostic Cohort among LETBI participants

### TES Criteria and Prevalence

*Head Trauma Exposure*. In addition to TBI, 138 (46.8%) participants had RHI exposure deemed to meet TES criteria (see Table 2).

**Table 2:**
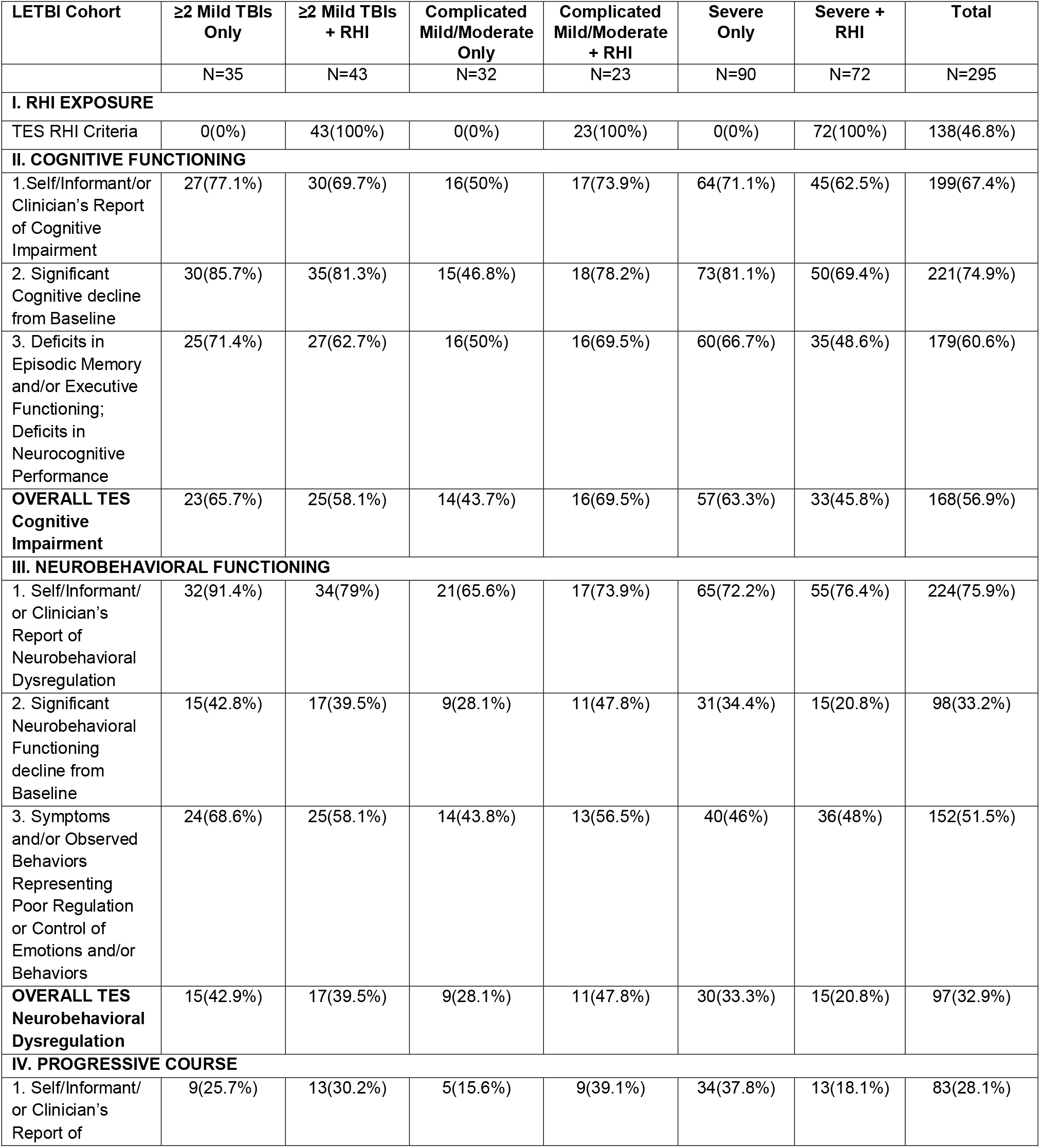
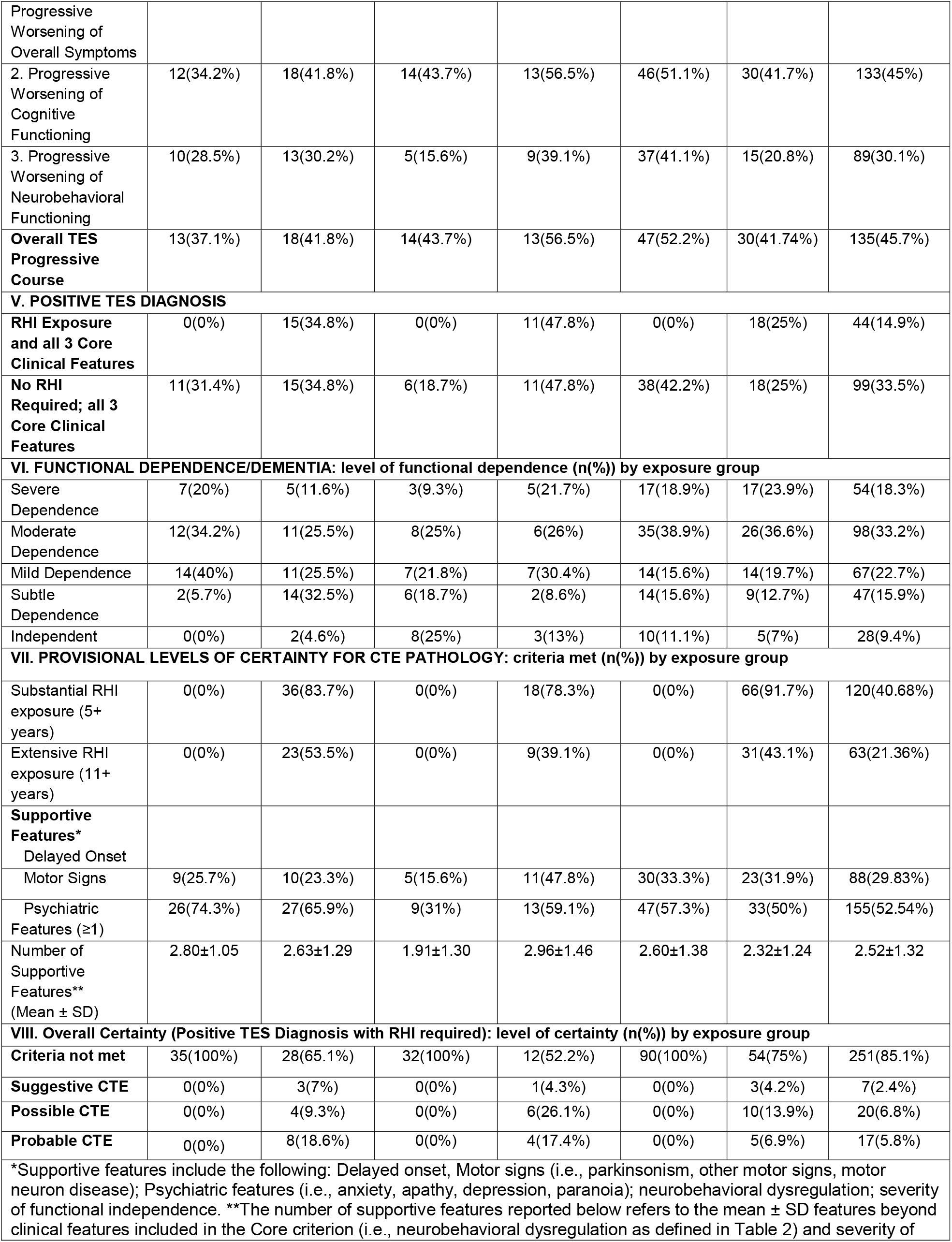
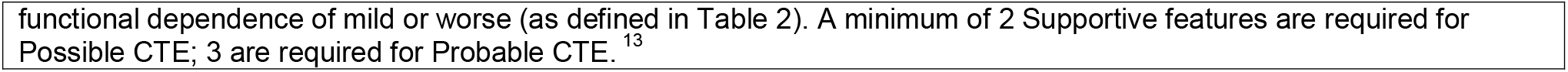
Proportion of sample meeting Core TES Criteria by head trauma exposure group.

#### Core Clinical Features

Overall, 56.9% of the sample met the TES core criterion of cognitive impairment (Table 2). A greater proportion of individuals with moderate TBI+RHI had cognitive impairment compared to those with isolated moderate TBI (69.6% versus 43.8%, respectively) while a lower proportion of those with severe TBI+RHI had cognitive impairment than those with severe TBI alone (45.8% and 63.3%, respectively). Rates were similar in the mild TBI groups with and without RHI.

One third of the full sample (32.9%) met TES criteria for overall neurobehavioral dysregulation. Neurobehavioral symptoms were reported by self or informant by an overwhelming majority of the sample (75.9%), and one third of the sample showed neurobehavioral decline from baseline (33.2%). We found that half (51.5%) had poor regulation/control of symptoms or behaviors. The rates of meeting each criterion across groups ranged from 20.0-47.8% overall; the mild group without RHI had rates consistently greater than 40%, and as high as 68% on all neurobehavioral criteria.

In the total sample, 45.7% of participants met criteria for progressive course of clinical features, with little variability among the TBI severity groups. When separately examining progressive course of cognitive and neurobehavioral features, the former is found to be far more common across all exposure groups (see Table 2). Most individuals with cognitive decline also had neurobehavioral worsening.

#### Level of Functional Dependence/Dementia

Only one quarter of participants met criteria for either functional independence (9.4%) or subtle dependence (15.9%). Participants with mild TBI without RHI showed a surprisingly high level of severe dependence (20.0%) compared to mild TBI with RHI (11.6%), and moderate TBI without RHI (9.3%); however the other groups showed comparable rates of severe dependence (18.9-23.9%)

#### Prevalence of Primary Diagnostic Criteria for TES

The Katz et al. ^13^ TES diagnostic criteria require RHI exposure and all three core clinical criteria; 14.9% of the LETBI sample met TES criteria (Table 2). We explored the prevalence of clinical TES features among those with isolated TBI by lifting the RHI exposure criterion, and found that 33.5% of the LETBI sample met all core clinical criteria. We tested whether there were significant differences between individuals with and without RHI exposure on key criteria, including cognitive impairment (χ^2^ (1, 295) = 1.17, p = 0.291), neurobehavioral dysregulation (χ^2^ (1, 295) = 0.35, p = 0.620), progressive course (χ^2^ (1, 295) = 0.25, p = 0.641), and overall TES criteria (χ^2^ (1, 295) = 0.33, p = 0.622). We found no significant differences.

#### Provisional Level of Certainty for CTE Pathology

RHI is required for even “suggestive” certainty of CTE, as isolated TBI does not meet the head trauma exposure criteria. In this sample of individuals with TBI, rates of suggestive, possible, and probable were 2.4%, 6.8%, and 5.8%, respectively (see Table 2). When we examined specific criteria, we found that participants with RHI overwhelming met criteria for Substantial RHI (5+ years of exposure), ranging from 78.5-88.0%, and a substantial portion met criteria for Extensive (11+ years; 39.1-53.5%). Just over one quarter of the full sample had motor signs, and there were no consistent patterns by TBI severity or RHI. Over half of the full sample endorsed at least one psychiatric feature (52.5%), again the rates did not differ by TBI or RHI in a discernible pattern.

### Predictors of TES

We found that no injury or demographic variables significantly predicted the likelihood of meeting all 3 Core Clinical Criteria for TES (Table 3). When we limit the sample to those who met TES RHI exposure criteria (n=138), being married (compared to being single) was weakly associated with the likelihood of meeting TES Core Criteria (OR=3.02, p=0.10); this association did not achieve statistical significance. No other predictors were statistically significant.

**Table 3:**
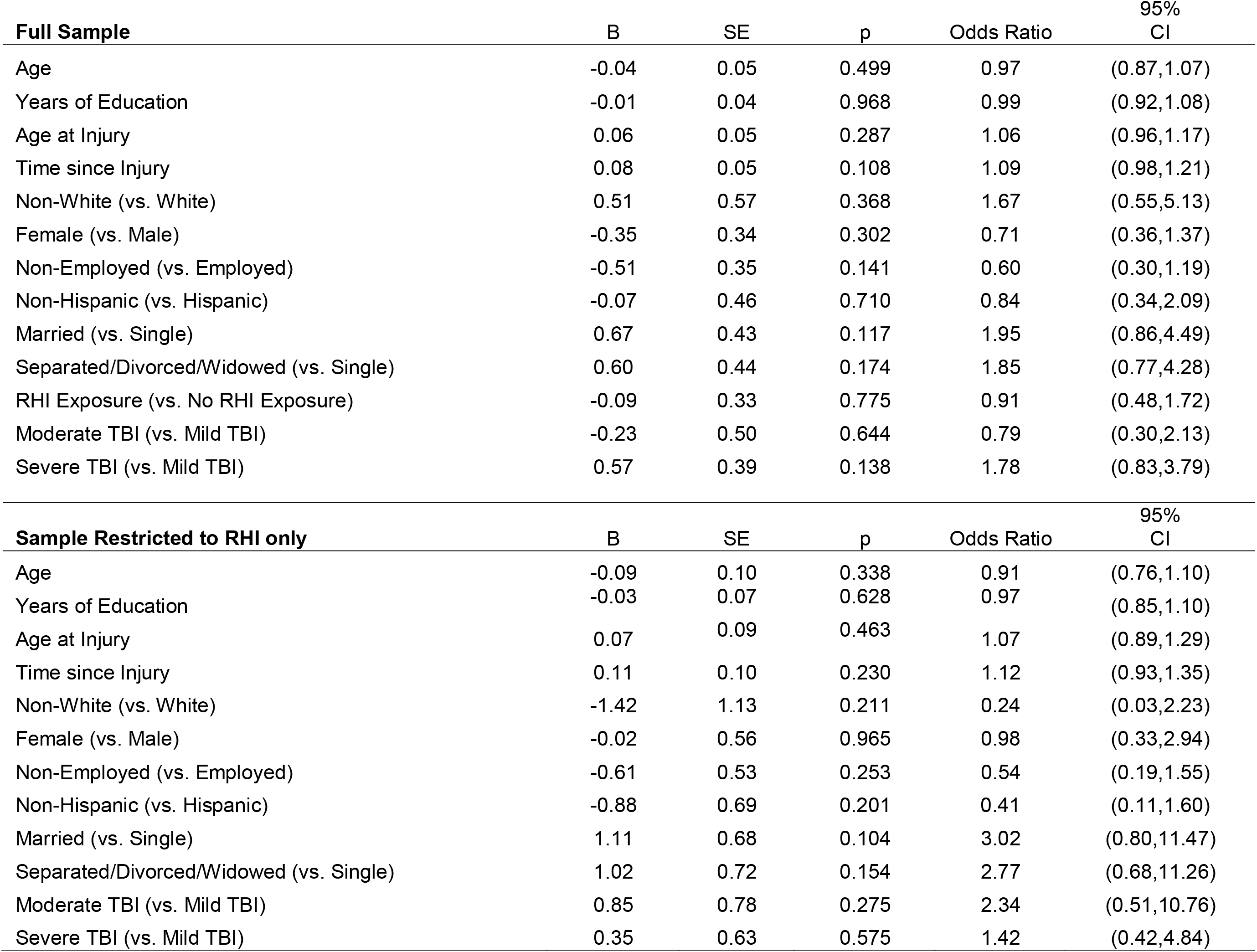
Summary Model of Selected Candidate Predictors for TES Diagnosis.

### Disease Comorbidity across groups defined by RHI exposure and TES per core criteria

Comorbid medical conditions are presented for descriptive purposes in Supplementary Table S4. Those meeting TES core clinical criteria and RHI exposure criteria had the highest rates of ischemic attacks (7.5%), migraine (46.3%), joint disease (41.8%), behavioral health conditions (18.2%), hearing disorders (2.3%), infectious diseases (4.5%), metabolic disorders (2.3%), and other neurological conditions (9.1%). Rates of epilepsy (14.1%), sleep apnea (23.9%) hypertension (21.7%), cancer (9.8%), coronary disease (16.7%), and respiratory disease (25.9%) were greatest among those who met TES core clinical criteria without RHI exposure.

## DISCUSSION

We applied the Katz et al^13^ TES diagnostic criteria using prospectively collected data in the LETBI cohort of individuals with isolated TBI, of whom 46.8% also had RHI exposure. Only those with RHI exposure were eligible for TES diagnosis, 14.9% met diagnostic criteria. When considering core clinical features irrespective of RHI exposure, a majority of our cohort met core TES criteria for cognitive impairment (56.9%) and progressive course (45.7%), while 32.9% met core criteria for neurobehavioral dysregulation. A greater proportion of the LETBI cohort met criteria for all 3 core clinical features of TES (Cognitive Impairment, Neurobehavioral Dysregulation, and Progressive Course) if they had isolated TBI compared to those with RHI+TBI. There were no injury or demographic features that predicted likelihood of meeting the core clinical criteria. Those who met full TES diagnostic criteria (i.e., RHI exposure and all 3 TES core clinical criteria) had the greatest rates of comorbid health conditions. To our knowledge, this is the first study to use prospectively collected in-vivo data to document the rates of TES clinical features in a community-based sample of individuals with isolated TBI.

It has been previously established that the clinical features of TES are common in clinical and community samples, ^10,37^ but many prior studies were limited by lack of head trauma exposure data. Current findings indicate that the core clinical features of TES are highly prevalent among those with isolated TBI. Unlike prior studies, however, our findings in a large sample of individuals with TBI with and without RHI underscore the importance of RHI for in-vivo diagnostics of TES. CTE neuropathology is rarely seen in the brain tissue of decedents without RHI exposure^38-40^ or in cases of individuals with head trauma sustained in etiologies other than contact sports, ^41-43^ suggesting RHI may be a necessary but insufficient condition for the development of CTE neuropathology. When we interpret current findings in the context of extant postmortem studies, they support the TES Consensus panel decision to require RHI, not TBI alone, in the exposure criterion for TES. ^13^ To the extent that CTE neuropathology is unique to RHI, so too should TES criteria; TES may not be an appropriate diagnosis in those without RHI exposure. Our results suggest that isolated TBI that results in multi-domain clinical impairment may be one of the few clinical conditions that, per current TES criteria, ^13^ may be considered to “fully account” for all in-vivo diagnostic features of TES.

The current finding that RHI is so common in individuals with TBI suggests that TBI exposure should similarly be considered in studies of RHI and CTE to further understand the independent and interactive implications for clinical symptoms and pathological processes. Fifty-six percent of male former contact sport athletes presumed to have RHI met 2014 TES criteria^44^ and rates were highest (70%) among those with ≥4 TBIs. Forty-one percent of professional boxers and mixed martial artists had TES per 2019 criteria, ^13^ and more “knockouts” increased risk. ^45^ Careful review^4,46^ of postmortem neuropathology from early studies of boxers^47^ found CTE (per revised consensus definition^46^) in less than half of boxers^48^ whose symptoms defined this disease four decades ago. ^3, 49^ It is possible that number of sport- and non-sport-related isolated TBIs sustained in the context of contact sports may reflect (and therefore confound) degree of RHI exposure in ways that are not generalizable to unselected samples. In the current community based LETBI study sample, which did not selectively recruit former contact sport athletes, we find high rates of TES clinical features among those with RHI and TBI across injury severity. Rates were similar among those with 2+ mild TBIs, which is consistent with prior studies demonstrating elevated rates of chronic symptoms among those with multiple concussions. ^50^

That rates of meeting all core TES clinical features were highest among those with isolated TBI in the current sample suggests that chronic and sometimes progressive sequelae of TBI may overshadow symptoms that may not reflect clinical manifestation of CTE neuropathologic change. Failure to carefully measure both lifetime TBI and RHI may result in inaccurate attribution of TES symptoms to CTE neuropathology. ^51,52^

The need to further refine in-vivo TES diagnostic criteria was well recognized by the 2019 Consensus panel, as reflected by explicit recommendation for researchers to apply these preliminary criteria to existing datasets. ^13^ A recent paper^53^ used expert adjudication to apply the 2014 TES criteria^9^ to informant and medical record data and found high sensitivity (0.97) but low specificity (0.21) to CTE neuropathology. ^5^ Cognitive symptoms were particularly predictive of CTE neuropathology. ^53^ Greater duration of contact sport participation (a proxy for RHI exposure) has been found to be directly related to CTE pathological burden, which appears to be associated with symptom severity, at least in symptomatic samples. ^6-9,54^ If RHI exposure is a key factor that uniquely distinguishes CTE from other pathological processes that contribute to otherwise-indistinguishable clinical symptoms, in-vivo differential diagnosis will require clearly defined and etiologically-specific exposure thresholds for men and women. This work must be conducted in samples with carefully characterized lifetime head trauma exposure, and must include those with and without TBI and RHI. Together with further refinement of TES core clinical criteria, this approach would serve to define exposure thresholds and specific clinical symptoms among those with head trauma who are likely to have CTE neuropathologic change.

Limitations of the current study warrant consideration. The LETBI sample does not currently include individuals with RHI exposure or isolated TBI alone, precluding direct comparison of TES clinical features across distinct exposure etiologies. It should be noted that self-selection into a TBI research study may preferentially enroll those with more severe symptoms; this may at least partially explain the high rates of symptom burden observed even among those with ≥2 mild TBIs. Other sources of selection, such as the requirement that participants attend a 4-5 hour in-person study visit, may also exclude those with severe disability that precludes travel and study visit completion. The LETBI study began before consensus-based TES criteria had been proposed, and study measures were not selected to align with TES criteria. This precluded operationalization of all supportive TES features^13^ (e.g., delayed onset of symptoms) without using measures used to define other criteria (e.g., progressive course). Duration of RHI sustained in the context of military service and partner violence were not available for the full sample. Incomplete longitudinal data at the time of analyses required definition of change over time per methods described by Katz. ^13^ Small cell sizes across TBI severity and RHI groups requires cautions interpretation of observed differences. Decisions regarding how to use available data to operationalize TES criteria were subject to clinical judgment, and may vary across studies and measures. Finally, absent postmortem autopsy data, it is not possible to directly investigate the sensitivity of TES criteria to CTE neuropathology at this time.

To our knowledge, this is the first study to apply the consensus-based TES research criteria in a community-based sample of well-characterized participants with a wide range of head trauma exposure histories. When interpreted alongside a growing body of postmortem studies suggesting CTE to be unique to RHI, current findings underscore the potential centrality of RHI as a key exposure criteria for TES diagnosis intended to facilitate in-vivo diagnosis of clinical symptoms reflective of CTE neuropathologic change. There is an urgent need to identify distinct clinical features most closely associated with CTE neuropathologic change so that those at risk for this distinct pathological process can be identified during life. Clinical and biological markers that can be quantified and monitored during life would inform clinical trial selection to facilitate inclusion of individuals whose symptoms are likely driven by the same underlying pathological process. More detailed understanding of RHI exposure thresholds will inform risk tolerance thresholds to allow athletes and others to make informed decisions about RHI exposure limits. As efforts to improve in vivo diagnostics progress, practicing clinicians should use available pharmaceutical and neurobehavioral interventions to address primary symptoms, recognizing that they may not share a common underlying pathophysiology.

## Data Availability

All data produced in the present work are available upon reasonable request to the authors and to the National Institute of Health's Federal Interagency Traumatic Brain Injury Research Repository.

