## Supplemental Tables for "Traumatic Encephalopathy Syndrome in the Late Effects of Traumatic Brain Injury (LETBI) study cohort"

**Supplemental Table S1:** TES Criteria and Method of Ascertainment and Operationalization within LETBI Study

| **TES Criteria Component** | **LETBI Methods of Ascertainment** | **LETBI Operationalization** |
| --- | --- | --- |
| **I. Substantial Exposure to Repetitive Head Impacts: at least 1 of the following is required** | | |
| A. High-exposure contact or collision sports   ≥5 years American football, at least 2 of which at high school level or beyond  Substantial years (e.g., ≥5) of other organized contact/collision sports | Review of available medical or public records to confirm head injury and severity of injury.  Self-reported exposure to contact/collision sports, military service or other sources of repetitive head impact on the Brain Injury Screening Questionnaire (BISQ). ^26^  Informant-reported exposure to contact/collision sports, military service or other sources of repetitive head impact on the BISQ. ^26^ | 1. At least five years of cumulative football play with at least two years occurring after the age of 13  2. At least five years cumulative play of collision sports or repeated blows to the head from sports from BISQ^26^ |
| B. Military service   Combat exposure to multiple blast/explosions  Noncombat exposure to explosions  Multiple blows to the head over time |  | 3. At least two years’ combined exposure to combat training and/or deployment  or  4. At least two years of military combat |
| C. Other sources of RHI   Multiple head impacts over an extended period of time (e.g., intimate partner violence) |  | 4. Report of at least two incidents of intimate partner violence resulting in injuries to the head or neck (e.g., nonfatal strangulation) and at least 2 years in a violent relationship. |
| **II. Core Clinical Feature** | | |
| **A. Cognitive Impairment: ALL 4 are required** | | |
| Self-, Informant-, or Clinician- report of cognitive impairment | Self-report question, “Compared to other people your age, how would you rate your memory?”  or  National Alzheimer’s Coordinating Center (NACC) Clinician Judgment of Symptoms items^29,31^ indicating difficulty with:  “Memory, Orientation, Executive functioning, Language, Visuospatial function, Attention/concentration, Fluctuating cognition” | 1. Response of either ‘fair’ or ‘poor’ on the Midlife in the United States (MIDUS) self-report memory question, ^30^  or  2. Affirmative response (‘Yes’) on any National Alzheimer’s Coordinating Center (NACC Clinician Judgement of Symptoms items 4a-h. ^29,31^ |
| Significant decline from baseline functioning | NACC Clinician Judgment of Symptoms item 1, ^29,31^ “Does the subject report a decline in memory (relative to previously attained abilities)?”  Pre-Morbid IQ Calculation: Age, Sex, Education. No job occupation variable available for IQ estimation, substituted a value of ‘5’ for all individuals. ^55^ Wide Range Achievement Test – 4 (WRAT-)4^56^ | 1. ‘Yes’ response to NACC Clinician Judgement of Symptoms items 1, 2, or 4a-h. ^29,31^ Endorsing any response besides ‘assessed previously’ NACC Clinician Judgement of Symptoms #5^29,31^  or  2. Pre-morbid IQ function was based on Age and Education Years. FSIQ=((0.017)(Age)-(1.53)(1=All sexes)- 11.33(1=All Races) +2.97(Education Years)+ 1.01(5 for all occupations)+74.05. ^33,34^ A t-score of at least 1.5 standard deviations below estimated pre-morbid IQ on any neuropsychological assessment. ^57,58^ |
| Deficits in episodic memory and/or executive function | Deficits in memory/executive function from the NACC Clinician Judgement of Symptoms^29^ included ‘Yes’ responses to the following cognitive impairment prompts with difficulty in: Memory, Orientation, Executive functioning, Language, Attention/concentration.  Comprehensive standardized neuropsychological battery assessing executive functioning, memory, estimate of premorbid cognitive functioning, including:  1. California Verbal Learning Test (CVLT-II) Short Form^59^  2. Wechsler Memory Scale – Logic Memory (WMS-IV) ^60^  3. Wechsler Adult Intelligence Scale – Digit Span, Coding, Symbol Search (WAIS-IV) ^61^ 4. Trails A&B^62^  5. Controlled Oral Word Association Test (COWAT) ^63^  6. Animal Fluency Test^64^  7. Rey Complex Figure Test^65,66^ | 1. Deficits in memory/executive functioning defined as either 1) ‘Yes’ response to any NACC Clinician Judgement of Symptoms item 4a-h, ^29^  or  2. A t-score ≤35 on any neuropsychological assessment 1-7 ^+, 57,58^ |
| Substantially impaired performance on formal neuropsychological testing (if available) |  | 1. A t-score ≤35 on any neuropsychological assessment 1-7 ^57,58^ |
| **B. Neurobehavioral Dysregulation: ALL 3 are required** | | |
| Self-, informant-, or clinician-reported neurobehavioral dysregulation | Self- and clinician- administered assessments:  1. NACC Clinician Judgment of Symptoms items^29,31^ referring to ‘any kind of behavioral symptoms’ including, disinhibition, irritability, agitation, personality change (bizarre behavior or behavior uncharacteristic of the subject), or other behavioral problems,  or  2. Barratt Impulsivity Scale II^67^ (BIS) including:  a) ‘Always or Often’ responses on ‘do things without thinking,’ ‘say things without thinking,’ ‘act on impulse,’ ‘act on the spur of the moment,’ ‘buy things on impulse,’ or ‘spend more than I earn’ OR  b) ‘Rarely/Never’ responses on ‘I am self-controlled’  or  3. Brown-Goodwin Lifetime History of Aggression^68^ affirming responses during adulthood including ‘angry outbursts/temper tantrums,’ ‘severe arguments with others,’ ‘physical fights,’ ‘destroyed someone’s/own property,’ or ‘done anything against the law without getting caught’ | 1. ‘Yes’ response to any NACC Clinician Judgment of Symptoms item 8 or 9d-g, ^29,31^  or  2. A t-score ≤35 for the reverse-scored BIS, ^67^  or  3. Any responses of ‘Always’ or ‘Often’ for BIS items 2, 12, 15, 18, 21, 25, ^67^ any response of ‘Rarely/Never’  or  ‘Occasionally’ for BIS item 6, ^67^  or  4. Any Brown Goodwin Lifetime History of Aggression^68^ response of ‘Occasionally’ or ‘Often’ during adulthood for items 3, 5, 6, 7, 8 if ‘drug dealing’ or ‘racing,’ 9, 10, 11, or 12. |
| Significant change from baseline functioning |  | 1. ‘Yes’ response to any NACC Clinician Judgment of Symptoms item 9d-9g, ^29,31^ or  2. A t-score decline of at least 1.5 standard deviations between study visits on any neurobehavioral assessment. |
| Poor regulation/control of emotions or behavior (Explosiveness, Impulsivity, Rage, Violent Outbursts, Mood swings/emotional lability) |  | 1. ‘Yes’, to any NACC Clinician Judgement of Symptoms item 8 or 9d-g^29,31^ or  2. A t-score ≤35 for BIS^57,58,67^ or  3. Any Brown-Goodwin Lifetime History of Aggression^68^ response of ‘Occasionally’ or ‘Often’ during adulthood for items 3, 5, 6, 7, 8 if ‘drug dealing’ or ‘racing’, 9, 10, 11, or 12 |
| **C. Progressive Course** | | |
| Evidence of progressive worsening of clinical features | Self-, informant-, or clinician-reported decline, per: 1) NACC Clinician Judgement of Symptoms, ^29,31^ 2) MIDUS Self Rated Health Questionnaire, ^30^ 3) RAND Item Short Form Health Survey (SF-36) ^69^  Calculated difference in scores on standardized neuropsychological tests between time points. Progressive course calculated separately for cognitive and behavioral domains. | 1) Response of either ‘Gradually Progressive’ or ‘Stepwise’ to NACC Clinician Judgement of Symptoms item 20^29,31^ assessing overall course of the disease.  2) MIDUS Self-reported memory^30^ decline of at least 1-unit (i.e. fair to poor) on the Likert Scale, or  3) For those with at least two study visits, cognitive decline was defined as 1.5 standard deviation decline on any neurocognitive assessment. Behavioral decline was defined as a) 1.5 standard deviation decline on any neurobehavioral assessment, or b)  1.5 standard deviation decline on either the RAND SF-36^69^ ‘Role Limitations due to Emotional Problems’ or the ‘Emotional Well-Being’ subscales. |
| **III. Not Fully Accounted for by Other Disorders (see Table 5)** | | |
| **D. Functional Dependence/Dementia** | | |
| Level of functional dependence based on impact of cognitive impairment and/or neurobehavioral dysregulation. Categorized as Independent, Subtle/Mild Functional Limitation, Mild Dementia, Moderate Dementia, Severe Dementia | Self-, informant-, or clinician-reported cognitive and social functioning, per 1) NACC Clinical Dementia Rating (CDR), ^29,31^ 2) Glasgow Outcome Scale-Extended, ^70^ 3) Neuro Quality of Life (NeuroQOL)^71^ social participation 4) NACC Functional Assessment Scale (FAS) ^29,31^ | 1. NACC CDR Global score 0=independent, 0.5=subtle, 1=mild, 2=moderate, 3=severe  2. GOS-E 3/4=severe, 5=moderate, 6=mild, 7=subtle, 8=independent  3. NeuroQOL “always” to all=independent, “often” to any=subtle, “sometimes” to any=mild, “rarely” to any=moderate, “never” to any=severe  4. FAS “normal”=independent, “has difficulty but does by self”=subtle, “requires assistance”=moderate, “dependent”=severe |
| CTE : Chronic Traumatic Encephalopathy; LETBI : Late Effects of Traumatic Brain Injury Project; TES: Traumatic Encephalopathy Syndrome | | |

**Supplemental Table S2:** Criteria and Supportive Features Used in Determining Provisional Levels of Certainty for CTE Pathology

| **CTE Provisional Certainty Component** | **LETBI Operationalization** |
| --- | --- |
| ***1. Suggestive of CTE:*** Meets TES criteria but does not meet the additional criteria for possible, probable, or definite CTE below. Suggestive of CTE pathology but does not cross the threshold for possible or probable CTE | |
| No additional criteria | N/A |
| ***2. Possible CTE:*** Meets TES criteria and must meet the first 2 criteria and a minimum of 2 supportive features | |
| A. Substantial exposure to a contact/collision sport: exposure to American football ≥5 y (including at least 2 at or beyond the high school level) or other contact/collision sport (e.g., boxing) to an equivalent extent | See Table 1 Section 1A-C |
| B. Cognitive impairment as defined in Table 1 | See Table 1 Section IIA |
| C. Plus a minimum of 2 supportive features as defined below, with Severity of Functional Dependence | Functional Dependence of subtle/mild functional limitation or worse |
| ***3. Probable CTE:*** Meets TES criteria and must meet the first 2 criteria below and a minimum of 3 supportive features | |
| A. Extensive exposure to a contact/collision sport: exposure to American football ≥11 y (including at least some at college level); or boxing or other sports with high-level exposure to RHIs to an equivalent extent^c^ | At least eleven years of cumulative football play (some at college level) or other contact collision sport, or at least eleven years of military combat exposure or non-combat repetitive head impacts during military service/training, or at least eleven years of intimate partner violence resulting in injuries to the head or neck |
| B. Cognitive impairment as defined in Table 1 | See Table 1 Section IIA |
| C. Plus a minimum of 3 supportive features as defined below, with Severity of Functional Dependence | Functional Dependence of mild dementia or worse |
| ***4. Definite CTE with TES:*** Meets TES criteria as well as CTE, confirmed by postmortem neuropathologic diagnosis based on current *National Institute for Neurological Disorders and Stroke* (NINDS )criteria for neuropathologic diagnosis of CTE. This is the gold standard for defining CTE disease entity. | |
| A. Postmortem neuropathologic diagnosis | N/A |
| ***Supportive Features*** | |
| 1. Delayed Onset Core clinical features begin following a clearly established period of stable functioning after the RHI exposure ends. | *Decline from baseline - no other information about delayed onset available*  See Table 1 Section IIA |
| 2. Motor Signs Parkinsonism | 1. Unified Parkinson’s Disease Rating Scale (UPDRS)^72^ Bradykinesia, Gait, Rigidity, and Tremor components scores ≥2  2. National Alzheimer’s Coordinating Center (NACC) Clinician Judgement of Symptoms^29,31^ item 14a gait disorder, item 14c tremor, item 17 Parkinsonism 3. Diagnosis of Parkinsons Disease |
| *Other motor signs* | 1. UPDRS^72^ item assessing Dysarthria ≥2 2. Balance score: mean z scores of Adult Changes in Thought Study Form 85 (ACT85)^73^ item side by side stand, semi-tandem stand, full-tandem stand, and UPDRS^72^ item postural stability, z≤-1.5 |
| *Motor neuron disease* | 1) Comorbid conditions under Other Neurological Disease: Autonomic Neuropathy, Trigeminal Neuralgia, Unspecified Idiopathic Peripheral Neuropathy, Hereditary Progressive Muscular Dystrophy, Other Convulsions, Dizziness and Vertigo. 2) Weakness: grip strength (per hand dynamometer) z score average of dominant and nondominant hands, z≤-1.5 3) NACC Clinician Judgement of Symptoms^29^ item 18 assessing amyotrophic lateral sclerosis |
| 3. Psychiatric Features Anxiety | 1) Neuro Quality of Life (NeuroQOL) Anxiety^71^ Tscore ≥65 2) Comorbid Anxiety disorder 3) NACC Clinician Judgement of Symptoms item 9i anxiety |
| *Apathy* | 1) NeuroQOL^71^ Depression item 8: "I felt that nothing was interesting" ≥3 2) NPI^74^ item 8 apathy/indifference ≥2 3) NACC Clinician Judgement of Symptoms^29^ item 9a apathy/withdrawal |
| *Depression* | 1) NeuroQOL^71^ Depression Tscore ≥65 2) Comorbid Depressive disorder 3) NACC Clinician Judgement of Symptoms^29^ item 9a assessing Depressed mood |
| *Paranoia* | 1) Comorbid conditions: Schizoaffective Disorder, Unspecified Psychosis, Unspecified Dissociative Disorder 2) Neuropsychiatric Inventory (NPI)^74^ item 2 delusions, NPI item 3 hallucinations 3) NACC Clinician Judgement of Symptoms^29^ item 9 psychosis-visual hallucinations; psychosis-auditory hallucinations; psychosis-abnormal, false, or delusional beliefs |
| CTE: Chronic Traumatic Encephalopathy; LETBI : Late Effects of Traumatic Brain Injury Project; TES: Traumatic Encephalopathy Syndrome | |

**Supplemental Table S3:** RHI Exposure Duration

|  |  |  | **Years of Exposure** | |
| --- | --- | --- | --- | --- |
| **Exposure** |  | **N** | **Range** | **Mean ± SD** |
| Football |  | 49 | 1-15 | 5.94±3.7 |
| Hockey |  | 11 | 3-45 | 16.27±13 |
| Lacrosse |  | 17 | 2-13 | 5.94±2.93 |
| Rugby |  | 8 | 2-6 | 3.88±1.46 |
| Soccer |  | 61 | 2-57 | 10.89±9.64 |
| MMA |  | 34 | 1-43 | 7.94±7.89 |
| Wrestling |  | 4 | 4-11 | 5.75±3.5 |
| Gymnastics |  | 5 | 4-8 | 6±1.87 |
| Cheerleading |  | 4 | 3-12 | 8.25±3.77 |
| Military |  | 26 | 0-23 | 4.77±4.85 |
| IPV |  | 32 | 0-54 | 22.26±14.69 |

**Supplemental Table S4:** Prevalence of Medical Comorbidities by head trauma exposure group

| **Medical Conditions** | **TES negative RHI negative** | **TES negative RHI positive** | **TES positive RHI negative** | **TES positive RHI positive** | **Total** |
| --- | --- | --- | --- | --- | --- |
|  | N=100 | N=96 | N=54 | N=45 | N=295 |
| Epilepsy | 4(4%) | 8(4.1%) | 10(18.5%) | 3(6.7%) | 25(8.5%) |
| Parkinson’s Disease | 1(1%) | 1(0.5%) | 0(0%) | 0(0%) | 2(0.7%) |
| Sleep Apnea | 15(15%) | 21(10.7%) | 13(24.1%) | 9(20%) | 58(19.7%) |
| Restless Leg Syndrome | 6(6%) | 7(3.6%) | 2(3.7%) | 0(0%) | 15(5.1%) |
| Hypertension | 19(19%) | 31(15.8%) | 12(22.2%) | 8(17.8%) | 70(23.7%) |
| Stroke or Cerebral Hemorrhage | 12(12%) | 10(5.1%) | 4(7.4%) | 4(8.9%) | 30(10.2%) |
| “Small Strokes”/transient ischemic attack | 0(0%) | 3(1.5%) | 0(0%) | 3(6.7%) | 6(2%) |
| Migraine | 22(22%) | 31(15.8%) | 12(22.2%) | 19(42.2%) | 84(28.5%) |
| Cancer | 9(9%) | 6(3.1%) | 8(14.8%) | 2(4.4%) | 25(8.5%) |
| Eye Disease or Injury | 27(27%) | 28(14.3%) | 16(29.6%) | 10(22.2%) | 81(27.5%) |
| Joint Diseases | 24(24%) | 29(14.8%) | 18(33.3%) | 21(46.7%) | 92(31.2%) |
| Digestive Problems | 25(25%) | 30(15.3%) | 11(20.4%) | 10(22.2%) | 76(25.8%) |
| Thyroid Disease | 14(14%) | 7(3.6%) | 7(13%) | 4(8.9%) | 32(10.8%) |
| Coronary Diseases | 4(4%) | 9(4.6%) | 9(16.7%) | 4(8.9%) | 26(8.8%) |
| Respiratory Diseases (including Pneumonia) | 11(11%) | 20(10.2%) | 14(25.9%) | 8(17.8%) | 53(18%) |
| Behavioral health conditions^1^ | 9(9%) | 14(7.1%) | 7(13%) | 8(17.8%) | 38(12.9%) |
| Hearing/Ear Disorders^2^ | 0(0%) | 0(0%) | 1(1.9%) | 1(2.2%) | 2(0.7%) |
| Infectious Diseases^3^ | 1(1%) | 0(0%) | 1(1.9%) | 2(4.4%) | 4(1.4%) |
| Metabolic Disorders^4^ | 3(3%) | 0(0%) | 0(0%) | 1(2.2%) | 4(1.4%) |
| Other Neurological Conditions^5^ | 3(3%) | 1(0.5%) | 2(3.7%) | 4(8.9%) | 10(3.4%) |
| Physical Injury^6^ | 1(1%) | 0(0%) | 1(1.9%) | 0(0%) | 2(0.7%) |
| Reproductive System/Genitourinary Diseases^7^ | 1(1%) | 2(1%) | 2(3.7%) | 0(0%) | 5(1.7%) |
| Other Diseases^8^ | 3(3%) | 0(0%) | 1(1.9%) | 1(2.2%) | 5(1.7%) |
| Additional conditions as per ICD-9 codes: 1 mental illness, depression, generalized anxiety disorder, adjustment disorder, post-traumatic stress disorder, bipolar disorder, dissociative identity disorder, borderline personality disorder.  2 tinnitus, vestibular impairment, hyperacusis. 3 Syphilis, Human Immunodeficiency Virus (HIV), Staphylococcus infection, Infectious mononucleosis, Lyme Disease, Leptospirosis. 4 Vitamin D Deficiency, Methylene Tetrahydrofolate Reductase Deficiency, Disaccharidase Deficiency,  Hyperlipidemia, Disordered Copper Metabolism, Hyposmolality, Anemia. 5 Autonomic Neuropathy, Trigeminal Neuralgia, Unspecified Idiopathic Peripheral Neuropathy, Hereditary Progressive Muscular Dystrophy, Other Convolusions, Dizziness and Vertigo. 6 Injury of Unspecified Nerve at Shoulder/Upper Arm Level, Chronic Low Back Pain, Individual in Car Accident. 7 Calculus of Kidney and Ureter, Unspecified Urethral Stricture, Submucous leiomyoma of uterus, Polycystic Ovarian Syndrome, Testicular Hypofunction, Prostate Specific Antigen/Hypertrophy, Endometriosis, Other Specified Genital Prolapse, Pilonidal Cyst. 8 Pituitary Dwarfism, Unspecified Hemmorrhoids, Hernia, Billiary Cirrhosis, Other Chronic Nonalcoholic Liver Disease Acute glomerulonephritis with Lesion, Renal Failure, Other Psoriasis, Disorders of Vestibular Function, Pituitary Adenoma. | | | | | |
